# Evidence for cross-reactivity and protection against Bundibugyo virus disease after heterologous vaccination: a systematic review and meta-analysis

**DOI:** 10.64898/2026.09.14.26362805

**Authors:** Eva Stadler, Mariah Csolle, Ainslie Mitchell, Ece Egilmezer, Karen M Elias, Bronwyn A Bailey, Deborah Cromer, Tari Turner, David S Khoury, Miles P Davenport

## Abstract

There are currently no approved vaccines or medical countermeasures for use in prevention or treatment of Bundibugyo virus disease. A critical question is whether the existing vaccines approved for use against Ebola virus disease could provide sufficient cross-reactivity and cross-protection against Bundibugyo virus disease. We performed a systematic review and meta-analysis of published studies of antibody cross-reactivity to Bundibugyo virus (BDBV) after vaccination with previously approved Ebola vaccines (Ervebo or Zabdeno/Mvabea). We find on average a 2.8-fold (95% predictive interval, PI: 2.6-3.0) drop in binding (n = 8 observations from 4 studies) and 3.1-fold drop (95% PI: 2.0-4.8) in neutralization (n = 2 observations from 1 study) between Ebola virus and BDBV antigens after Ervebo vaccination, which appears maintained over time after vaccination. Very limited data on cross reactivity after Zabdeno/Mvabea vaccination suggests a higher drop in recognition of BDBV (39.7-fold, 95% PI: 24.8-63.8; n = 3 observations from 2 studies). In addition to analysis of antibody recognition in humans, we also investigated evidence for vaccine protection from BDBV challenge in animal models after Ervebo or other recombinant vesicular stomatitis virus (rVSV) based vaccines containing only Ebola antigen. A single study in NHP showed non-significant vaccine protection from BDBV challenge, while a study in ferrets showed significant protection after both one and two doses. The available data demonstrate immune cross-reactivity and some evidence for protection in animals, although further clinical and pre-clinical data is urgently needed to inform decisions on the use of Ervebo to protect against Bundibugyo virus disease.

## Introduction

The current Bundibugyo Virus Disease (BVD) outbreak in the Democratic Republic of Congo was declared a Public Health Emergency of International Concern by the World Health Organization on 17 May 2026^1^. There is currently no vaccine or therapeutic approved specifically for BVD. However, there is an approved Ebola virus disease vaccine (Ervebo) that has been shown to be effective against Ebola Virus Disease (EVD)^2,3^. A major question is whether Ervebo can provide some degree of cross protection against BVD. There are currently no studies reporting clinical effectiveness of Ervebo against BVD, and such results are unlikely before 2027.

Given the rapidly evolving nature of the health emergency and response, and in the absence of studies reporting clinical protection in humans, here we undertook a systematic review and meta-analysis of other, less direct evidence (leveraging an existing evidence map^4,5^) that might inform health agencies of the potential utility of Ervebo against BVD. Firstly, we examined whether Ervebo (or the previously licensed Zabdeno/Mvabea vaccine) induces cross-reactive antibodies to Bundibugyo virus in humans. Although antibody binding or neutralization are not an established correlate of protection for Ebola infection, providing monoclonal antibody therapy to subjects testing positive by PCR (polymerase chain reaction) has been shown to be effective in preventing death in humans (although convalescent plasma was not associated with a significant improvement in survival from symptomatic infection)^6,7^. Further, antibody responses to Ebola glycoprotein have been observed to correlate with protection from death in animal studies of EVD^8–10^. Secondly, we examined whether Ervebo vaccination is associated with survival after Bundibugyo virus challenge in animal models of BVD. Animal data (non-human primate models of Ebola virus infection) have previously played a central role in supporting licensure of the Zabdeno/Mvabea EVD vaccine^11,12^.

Together these two analyses indicate provide evidence of cross-reactivity of Ervebo against Bundibugyo virus that may help inform decision-making on the use of Ervebo in the current Bundibugyo public health emergency.

## Methods

### Study identification and inclusion

To identify human and animal studies of the effects of Ervebo or Zabdeno/Mvabea vaccination, we screened articles from the exisiting living evidence map of Bundibugyo Virus (BDBV) studies, available at https://livingevidence.org.au/research-initiatives/about-feeva/evidence-maps/ (version: search date - 24 August 2026). Articles in the evidence map were independently screened in Covidence by two reviewers on title and abstract, and then full text, against pre-existing inclusion criteria. Differences are resolved through discussion and consultation with a third reviewer where required. Studies that meet inclusion criteria are independently tagged by two reviewers according to a structured classification scheme based on population, outcome and study type, and the results presented as an evidence map in EPPI-Mapper.

For assessment of the evidence of Bundibugyo virus immunogenicity after Ervebo or Zabdeno/Mvabea vaccination, we screened the evidence map for studies in humans that reported binding or neutralizing antibody responses to Bundibugyo virus after vaccination (specifically, filtering on Population = human, Intervention = vaccination, Outcomes = [Assay-Antibody binding OR Assay-Antibody neutralizing]). These publications were screened to identify studies that reported antibody responses to both Ebola virus and Bundibugyo virus after vaccination with either Ervebo or Zabdeno/Mvabea.

For assessment of clinical protection in animals after vaccination, we screened the BDBV evidence map for studies that reported survival after Bundibugyo virus challenge in vaccinated animals (specifically: Population = [Non-human primate OR mouse OR ferret OR guinea pig OR other animal], Intervention = infection-challenge, Outcome = survival). We then screened these publications for studies that reported survival outcomes after challenge with Bundibugyo virus of animals vaccinated with a recombinant vesicular stomatitis virus (rVSV)-based vaccine containing only Ebola virus antigens (compared with unvaccinated control animals). Because animal studies often used research-grade Ervebo rather than the licensed vaccine (or used different dosages to account for the animals’ different body weight), we included all rVSV vaccines containing only Ebola virus antigens in our analysis.

Studies from the evidence map were screened independently by two authors (ES, MC, DK, MPD). Conflicts were resolved by discussion and consensus. We also included studies identified through ongoing surveillance of the literature that met the inclusion criteria (Supplement p. 3 and p. 6).

### Risk of bias assessment

Risk of bias assessment was undertaken by two independent reviewers (MC/EE and TT for clinical studies and MC and EE for animal studies), with differences resolved through discussion, or consultation with a third reviewer where required.

For Bundibugyo immunogenicity in humans, included studies were assessed for risk of bias using the ROBINS-I V2 tool. This tool is designed to assess risk of bias in studies evaluating the impact of an intervention compared to another intervention or no intervention, without randomization. The ROBINS-I V2 tool is not entirely fit for purpose for assessing the risk of bias in studies assessing the cross-reactivity of vaccines for EVD to Bundibugyo virus. However, many of the concepts assessed are appropriate, and to our knowledge no risk of bias tool currently exists that better addresses the relevant issues.

For studies reporting survival after Bundibugyo virus challenge in vaccinated animals, included studies were assessed for risk of bias using the SYRCLE tool^13^, designed to assess risk of bias in intervention studies in animals.

### Data extraction

Data for Bundibugyo immunogenicity in humans were extracted from identified studies by one author (AM, SRK, EE, ES) and verified by another (ES, AM, DSK). Conflicts were resolved by discussion and consensus, with a third reviewer (MC) consulted if needed. We also contacted authors from all studies where supplementary data was not provided to request the raw data.

For assessment of survival of vaccinated animals challenged with Bundibugyo virus, data were extracted by one author (ES) and verified by another (AM).

### Data analysis

The aim of the analysis of Bundibugyo immunogenicity after Ervebo or Zabdeno/Mvabea vaccination was to assess the fold-drop in antibody binding or neutralization from Ebola virus to Bundibugyo virus. Thus, we used a mixed effects meta-regression to estimate a pooled fold-drop for each vaccine and antibody response (neutralizing or binding) combination and for each vaccine (regardless of antibody response). The outcome was the log_10_-fold-drop from Ebola to Bundibugyo virus. The model included a random intercept by study and was fitted to data using inverse variance weighting and the R-package ‘metafor’ (using the rma.mv function with restricted maximum likelihood method)^14^. Where the variance of the fold-difference between the Ebola and Bundibugyo virus antibody response was not provided, it was calculated from reported 95% confidence intervals, interquartile ranges, or raw data (Supplement p. 3). Data and model outputs (including 95% prediction intervals for the estimated fold-differences) were visualized as forest plots and publication bias was assessed using funnel plots. We report I^2^ with 95% confidence interval as a measure of between-study heterogeneity^15^. Residual heterogeneity (after accounting for moderators) was assessed using the Cochran Q (QE) test and indicated in the forest plot. We used an omnibus test (QM test) to assess whether moderators explain a significant amount of the variance in outcomes (also indicated in the forest plot). We tested two separate models with different moderators: one including vaccine as a moderator (Ervebo or Zabdeno/Mvabea, regardless of booster administration for Ervebo and dose-spacing for Zabdeno/Mvabea) and a second model including a combination of the antibody measure and the vaccine regimen (5 categories in total: neutralization and Ervebo, binding and Ervebo, binding and Ervebo with booster, binding and Zabdeno/Mvabea with 28-day spacing, and binding and Zabdeno/Mvabea with 56-day spacing).

Where antibody responses were reported to multiple Ebola virus strains, we used the response to Kikwit glycoprotein (GP) if possible (since the Ervebo vaccine contains the Kikwit GP antigen) and used responses to other Ebola virus strains in a sensitivity analysis (Supplementary Figures, p. 7). For studies reporting multiple Bundibugyo virus strains, we used the most recent Bundibugyo virus strain and compared Ebola and Bundibugyo virus fold-differences for all Bundibugyo virus strains to verify consistent results across fold-differences (Supplement 3).

Due to low or zero events in included data from animal models, we calculated 95% confidence intervals of the relative risk of death in vaccinated compared to control animals using the score method (using the R package ‘PropCIs’^16^). Since identified studies differed by animal model and number of vaccine doses, we did not perform a meta-regression or calculate a pooled estimate for this data.

For all statistical analyses, a significance level of 0.05 was used, i.e. p-value <0.05. All confidence and prediction intervals are 95% confidence and prediction intervals, and all statistical analyses were performed using R (version 4.5.2)^17^.

### Role of the funding source

The funder of the study had no role in study design, data collection, data analysis, data interpretation, or writing of the report.

## Results

### Bundibugyo virus immunogenicity after Ervebo or Zabdeno/Mvabea vaccination

We screened nine studies that were identified from either the Bundibugyo virus evidence map, citation searching, and on-going surveillance of the literature (**Figure S1**). Seven studies that reported immunogenicity in humans after vaccination with Ervebo or Zabdeno/Mvabea met our inclusion criteria^18–24^.

Risk of bias assessment using ROBINS-I V2 suggested that all included studies had potential for moderate, serious or critical risk of bias for potential confounding and missing data, and some studies also had potential for moderate or serious risk of bias in participant selection (**Figure 2**). In most cases this assessment is due to limited detail in the reporting of the characteristics of, or reasons for selection of, the study participants and samples. Without this information it is difficult to rule out the potential for bias in the study results due to confounding with factors including baseline seroreactivity or exposure to other factors that might influence immunogenicity during the studies. Similarly, limited reporting on reasons for selection of samples means we are unable to rule out the possibility that there are systematic reasons that the samples were included which might also be related to the outcome of interest. Nonetheless we chose to proceed with analyzing data from these studies, because we recognized that ROBINS-I is not an ideal tool for assessing such studies (discussed in more detail later). Further, given that assessment for risk of bias of this type of study is generally not conducted, it is important to note that these assessments do not necessarily indicate that the included studies had a higher risk of bias, or were of lower quality, than similar studies of this type^25–28^.

Data from 6 studies were included in the analysis (Smith et al.^24^ and Halbrook et al.^19^ reported data from the same study; we included data from Halbrook et al. only due to the larger sample size and separate reporting by location). The included data contained reports of antibody responses after Ervebo vaccination from 5 studies and after Zabdeno/Mvabea vaccination from 2 studies (1 study included both vaccines), neutralizing antibody responses from 1 study and binding antibody responses from 5 studies (**Figure 1**, **Table 1**).

**Figure 1.**
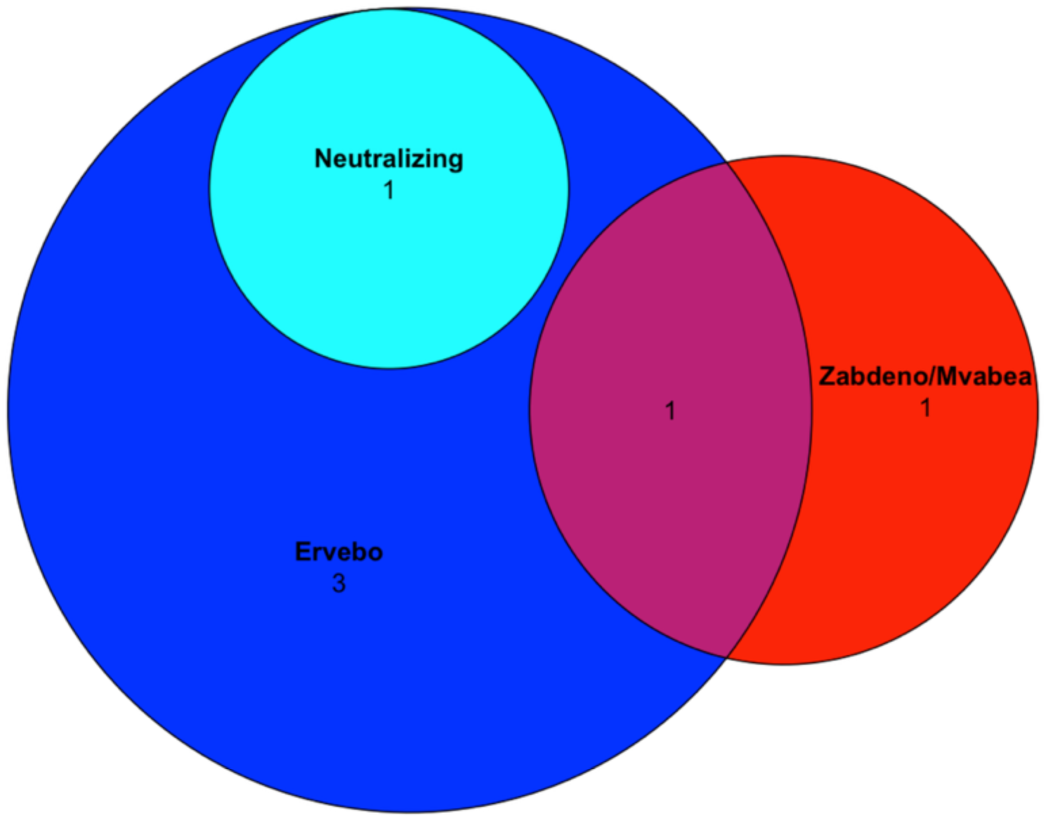
Venn diagram of identified studies included in the analysis. We included 6 studies in the analysis, 5 reporting antibody levels after Ervebo vaccination of which one reports neutralizing antibody responses and the remaining 4 report binding antibody responses. Two studies reported binding antibody responses after Zabdeno/Mvabea vaccination of which one includes both Ervebo and Zabdeno/Mvabea vaccination.

**Figure 2.**
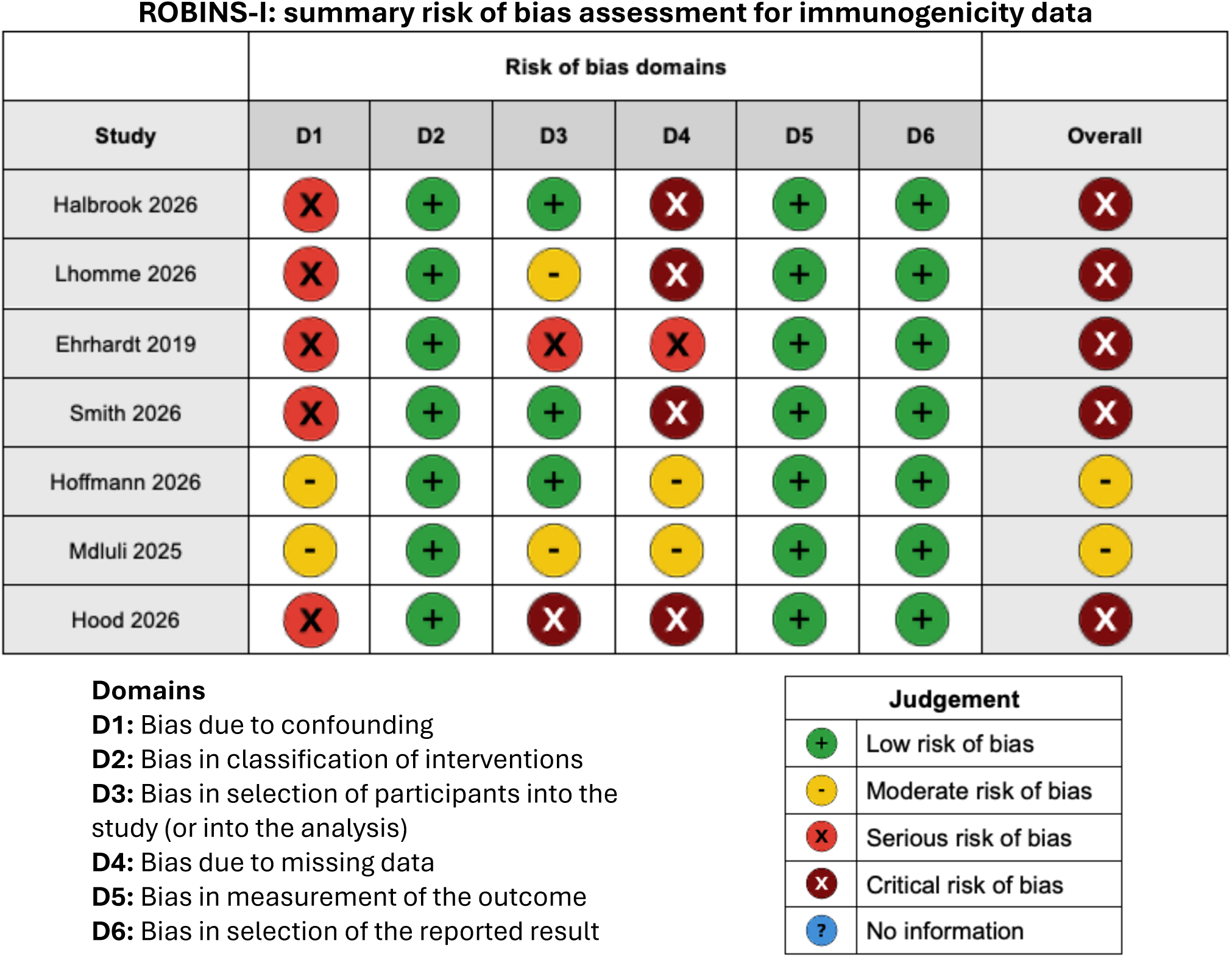
Summary of ROBINS-I risk of bias assessment for immunogenicity data.

**Table 1.**
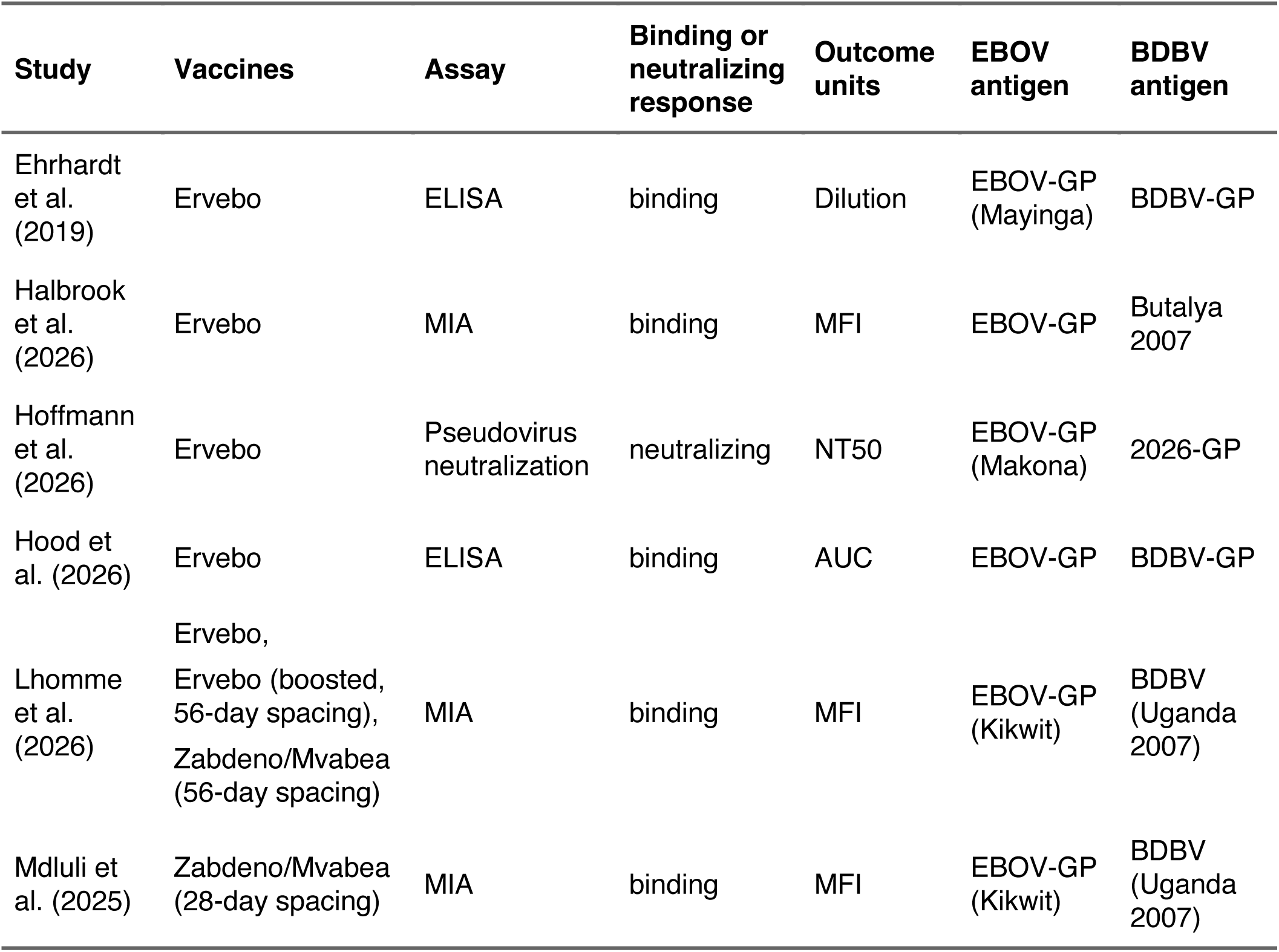
Bundibugyo and Ebola virus human immunogenicity studies overview. The data presented in this table was used in our main analysis. However, Hoffmann et al. (2026)^20^ also report neutralization titers for BDBV 2007/08-GP and 2012-GP, and Lhomme et. (2026)^22^ also report antibody binding to EBOV Mayinga GP (Supplement p. 3). Abbreviations: *AUC* area under the curve (OD vs dilution), *BDBV* Bundibugyo Virus, *EBOV* Ebola Virus, *ELISA* enzyme-linked immunosorbent assay, *GP* glycoprotein, *MFI* mean fluorescence intensity, *MIA* multiplex immunoassay, *NT50* 50% neutralizing titer.

| Study | Vaccines | Assay | Binding or neutralizing response | Outcome units | EBOV antigen | BDBV antigen |
| --- | --- | --- | --- | --- | --- | --- |
| Ehrhardt et al. (2019) | Ervebo | ELISA | binding | Dilution | EBOV-GP (Mayinga) | BDBV-GP |
| Halbrook et al. (2026) | Ervebo | MIA | binding | MFI | EBOV-GP | Butalya 2007 |
| Hoffmann et al. (2026) | Ervebo | Pseudovirus neutralization | neutralizing | NT50 | EBOV-GP (Makona) | 2026-GP |
| Hood et al. (2026) | Ervebo | ELISA | binding | AUC | EBOV-GP | BDBV-GP |
| Lhomme et al. (2026) | Ervebo, Ervebo (boosted, 56-day spacing), Zabdeno/Mvabea (56-day spacing) | MIA | binding | MFI | EBOV-GP (Kikwit) | BDBV (Uganda 2007) |
| Mdluli et al. (2025) | Zabdeno/Mvabea (28-day spacing) | MIA | binding | MFI | EBOV-GP (Kikwit) | BDBV (Uganda 2007) |

Reported data varied in the vaccine regimen (number of doses, dose spacing), assay and outcome units, Ebola virus antigen used, and Bundibugyo virus antigen used (**Table 1**). Nonetheless, we combined the fold-drops from Ebola to Bundibugyo virus antibody responses across studies. We found 3.12-fold (95% prediction interval, PI: 2.04-4.76) lower neutralizing antibody responses against Bundibugyo virus than against Ebola virus after a single-dose Ervebo vaccination (**Figure 3**). Binding antibody responses after vaccination with a single dose of Ervebo were 2.79-fold (95% PI: 2.58-3.02) lower to Bundibugyo virus antigen than to Ebola virus antigens (**Figure 3**). For all remaining vaccine regimens, there was only one observation each (Ervebo with a 56-day booster and Zabdeno/Mvabea with either a 28-day or a 56-day spacing between doses). Combined estimates per vaccine regardless of neutralizing or binding response, boosting, or dose spacing showed a 2.81-fold drop (95% PI: 2.60-3.03) for Ervebo and a 39.72-fold drop (95% PI: 24.75-63.76) for Zabdeno/Mvabea (**Figure S4**). A number of studies analyzed serum at different time intervals after vaccination (from 21 to 660 days), and there was no evidence of major changes in cross reactivity over time. The single study that reported on cross-reactivity after boosting with Ervebo had very wide confidence intervals, so it is unclear how boosting may affect cross-recognition of Bundibugyo virus. Here we used only one strain of Ebola virus for each study, which was the Kikwit where it was available, otherwise we used the strain that was available (**Table *1***). However, we noticed one study, Lhomme et al.^22^, had more than one strain (Kikwit, Mayinga). In a sensitivity analysis we found that using the Mayinga strain instead of Kikwit from this study did not impact the results (Supplement p. 3, **Figure S6** to **Figure S9**).

**Figure 3.**
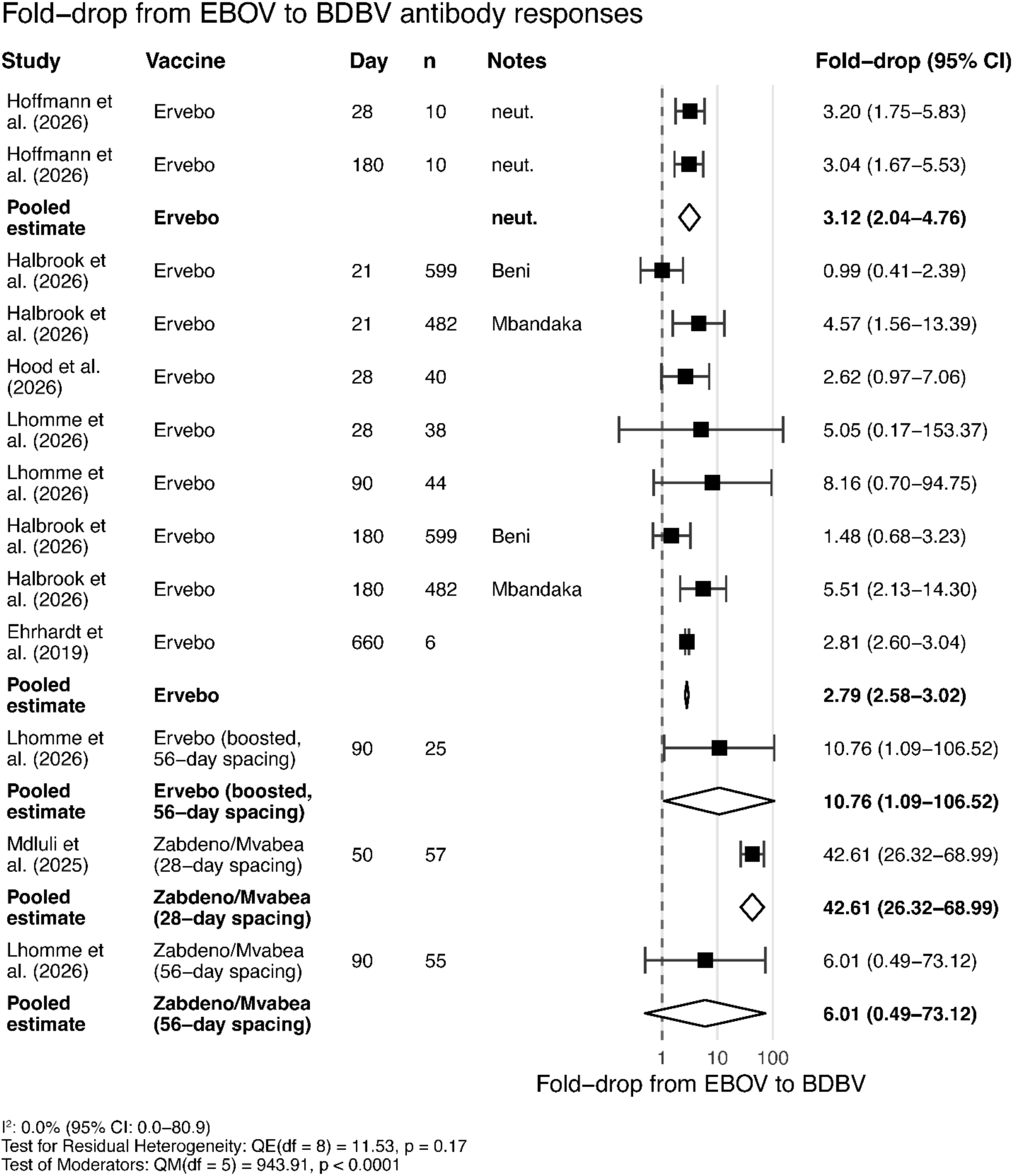
Fold-drop in Bundibugyo Virus (BDBV) antibody response compared to Ebola Virus (EBOV) antibody response. Data was grouped and a pooled estimate calculated by whether neutralizing (indicated by ‘neut.’ in the ‘Notes’ column) or binding antibodies (where ‘neut.’ is not indicated) were reported, and according to the vaccine used. Other entries in the ‘Notes’ column indicate the location from which samples were collected (where data from vaccination at different locations were reported in the same paper). For pooled estimates, we show 95% prediction intervals rather than confidence intervals (the reported data from each study are 95% confidence intervals). The associated funnel plot is shown in **Figure S3**. A sensitivity analysis using EBOV Mayinga instead of Kikwit in Lhomme et al. (2026)^22^ gave consistent pooled estimates for single dose Ervebo vaccination (**Figure S8**). Abbreviations: *BDBV* Bundibugyo Virus, *CI* confidence interval, *EBOV* Ebola Virus, *n* number of samples, *neut.* neutralizing antibody response.

Overall, we find a generally consistent fold-drop in Bundibugyo virus antibody responses compared to Ebola virus antibody responses after Ervebo vaccination, and very limited evidence suggesting a larger fold-drop in cross-reactivity between Ebola and Bundibugyo virus after Zabdeno/Mvabea vaccination.

### Survival after vaccination and Bundibugyo virus challenge in animal models

In the absence of clinical data on protection against BVD after vaccination with Ervebo in humans, we searched studies reporting survival after Bundibugyo virus challenge in animals vaccinated with Ervebo or another rVSV-based vaccine. To assess survival after Bundibugyo virus challenge, we included only vaccines that contain Ebola Virus (EBOV) glycoprotein antigens and no other antigens; Supplement p. 6, **Figure S2**). We identified 2 studies^29,30^ (**Table 2**) that reported vaccination of cynomolgus macaques and ferrets with rVSV-EBOV. One of the included studies (Wight et al. (2026)^30^) was published after the latest update of the Bundibugyo virus evidence map (24 August 2026). This study was identified through ongoing surveillance of the literature given the rapidly evolving nature of the BVD outbreak and was included as it met all pre-specified eligibility criteria.

**Table 2.** Overview of all identified studies that reported survival after Bundibugyo challenge in animals vaccinated with an rVSV-based vaccine including only Ebola virus antigens. Wight et al.^30^ did not further specify the vaccine used, but they state that rVSV-EBOV is also known by its tradename Ervebo, i.e. it is likely an rVSVΔG-ZEBOV-GP vaccine as in the study by Falzarano et al.^29^.

| Study | Animals | Number of animals | Vaccine | Days from initial vaccination to challenge | Challenge virus |
| --- | --- | --- | --- | --- | --- |
| Falzarano et al. (2011) | Cynomolgus macaques | 8 | rVSVΔG-ZEBOV-GP | 28 | Bundibugyo ebolavirus |
| Wight et al. (2026) | Ferrets | 16 | rVSV-EBOV | 28 | Bundibugyo ebolavirus (Butalya) |
|  | Ferrets | 16 | rVSV-EBOV (boosted, 14-day spacing) | 28 | Bundibugyo ebolavirus (Butalya) |

We assessed risk of bias using SYRCLE’s risk of bias tool for animal studies^13^, which identified that the two included studies had the potential for high risk of bias (**Figure 4**). For both studies, this judgement was driven by minimal reporting of randomization methods and allocation concealment (selection bias). For Falzarano et al. (2011)^29^, this was compounded by a lack of baseline group comparability and a disclosed conflict of interest, in which two senior authors hold intellectual property on the vaccine platform under evaluation (other bias). For Wight et al. (2026) ^30^, late addition of an unrandomized second control cohort (challenged 14 days after the first), introduced further risks of performance and detection bias.

**Figure 4.**
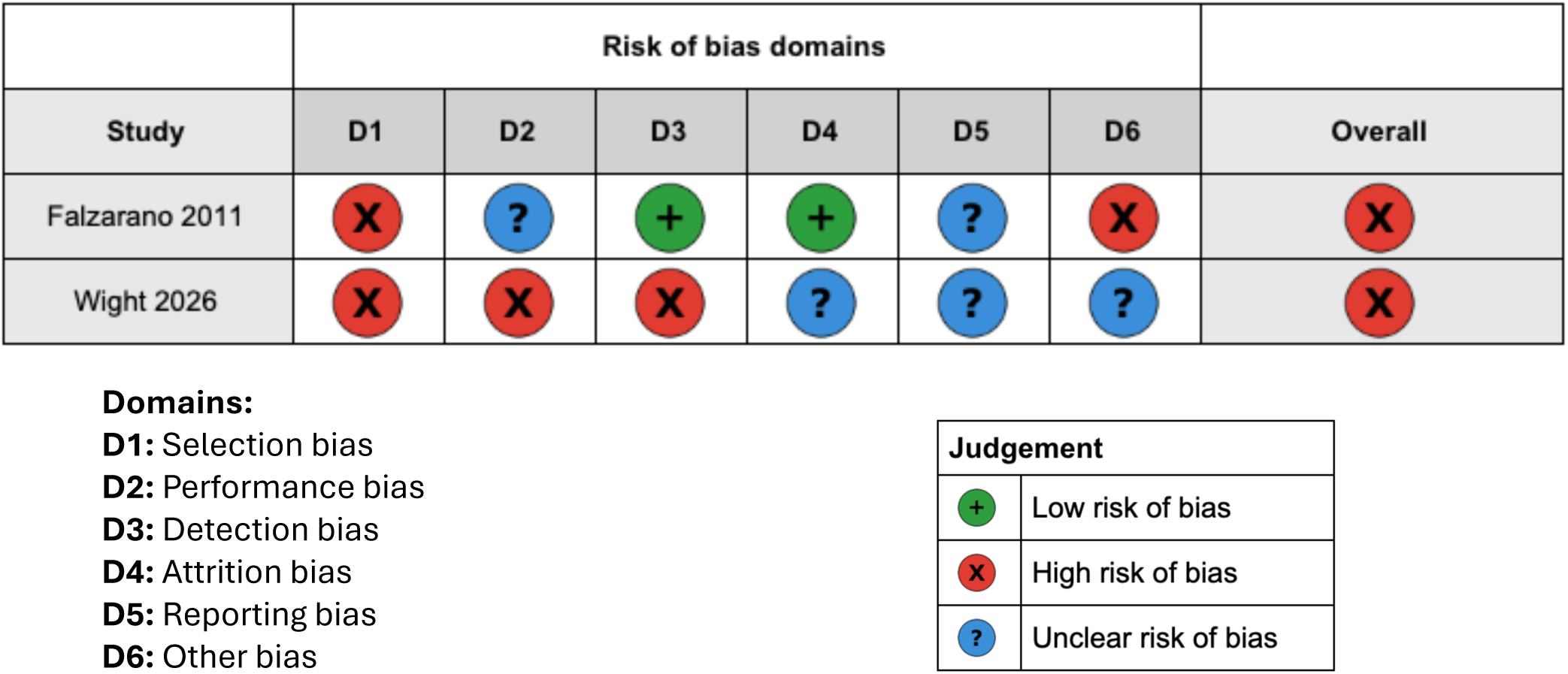
Summary of SYRCLE risk of bias assessment for animal studies.

All animals were challenged with Bundibugyo virus and survival was reported in vaccinated and unvaccinated control animals (**Figure 5**).

**Figure 5.**
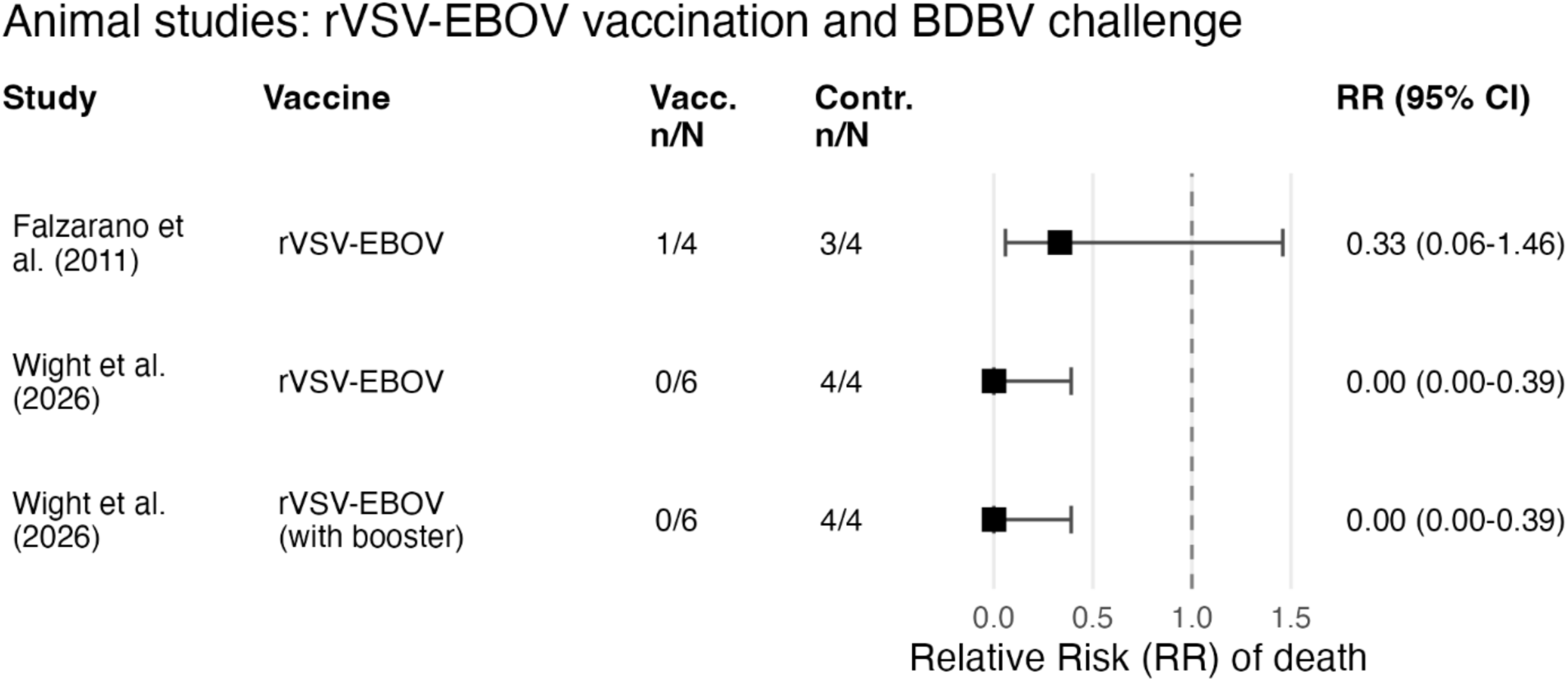
Relative Risk (RR) of death in vaccinated compared to control animals. RR < 1 indicates a lower risk for vaccinated compared to control animals, i.e. higher protection against death in vaccinated animals. Wight et al.^30^ included three different cohorts: single rVSV-EBOV vaccination (6 animals), rVSV-EBOV vaccination with a booster (6 animals), and a control cohort (4 animals) that we used to calculate RR and 95% CIs for both the single rVSV-EBOV and the boosted rVSV-EBOV cohorts. Abbreviations: *BDBV* Bundibugyo Virus, *CI* confidence interval, *contr.* control animals (not vaccinated), *n* number of events (deaths), *N* total number of animals, *RR* relative risk, *vacc.* vaccinated animals.

In the study by Falzarano et al., 3 of 4 unvaccinated cynomolgus macaques died after Bundibugyo virus challenge, compared to only 1 of 4 vaccinated animals. This indicated a trend for protection from death after vaccination (not statistically significant). In Wight et al. lethality of the Bundibugyo virus challenge in unvaccinated ferrets was 100% (4 of 4), whereas all ferrets vaccinated with either one dose (6 of 6) or two doses (6 of 6) of rVSV-EBOV vaccine survived Bundibugyo virus challenge. Overall, evidence of vaccine-induced protection against death after Bundibugyo virus challenge in animals is limited.

## Discussion

Vaccines or medical countermeasures are urgently needed to help address the 2026 Bundibugyo public health emergency in the DRC. To date there are no vaccines that have been shown to be effective in protecting against Bundibugyo virus disease, although there are a number in development. A major question is whether the existing vaccine approved for use against Ebola infection (Ervebo) might provide some degree of cross-reactivity and cross-protection from Bundibugyo Virus Disease (BVD). There are no clinical studies reporting BDV protection after Ervebo vaccination, but an observational study by LaRochelle et al.^31^ reported disease outcomes in a cohort of 37 hospital staff members early in the current BDV outbreak. The Africa CDC announcement on “The immediate implementation of the DRC Government’s decisions to stop the Bundibugyo Ebola emergency”^32^ also mentions observations from the DRC surveillance and the Mongbwalu cluster. However, to the best of our knowledge this observational evidence for potential protection against BVD after Ervebo vaccination has not yet been published. Thus, in the absence of data on the clinical efficacy of Ervebo against BVD, both the WHO and Africa CDC have cited individual studies of *in vitro* antibody responses to Bundibugyo virus as providing evidence of potential vaccine effectiveness. In addition, evidence of rVSV-EBOV vaccines protection in animal models has also been cited. To assess the totality of evidence for immune cross-reactivity and animal protection for Bundibugyo virus, we performed a systematic review to identify studies reporting antibody responses to Bundibugyo virus in humans after vaccination with Ervebo (or Zabdeno/Mvabea) and studies reporting survival of animals vaccinated with an rVSV-based vaccine and challenged with Bundibugyo virus.

Our analysis of the data on serological responses after Ervebo vaccination from 6 included studies showed a 2.79-fold (95% CI: 2.58-3.02) lower antibody binding to a Bundibugyo GP than to an Ebola GP (**Figure S4**). There was minimal heterogeneity between studies, and the result was similar between binding and neutralizing antibody responses. The meta-analysis presented here has several limitations. Most importantly, the current BVD research literature is limited with few studies reporting relevant data on Bundibugyo immunogenicity or clinical outcomes. Secondly, our analysis of risk-of-bias in these studies suggested there are important risks of bias in these studies. The ROBINS-I tool used to assess risk-of-bias is not entirely fit for purpose for the assessment of studies comparing *in vitro* assays of immune responses, and many of the concerns of potential for bias arose from limited reporting of reasons for selection of a subset of participants from a larger sample pool. This is the first known transparent assessment of risk of bias in this research area, and the use of ROBINS-I may overestimate risk of bias (meaning our assessment is conservative). However, the consistency of the drop in titer across multiple studies provides some reassurance in the robustness of the results. The accuracy and utility of risk of bias assessment of future immunogenicity studies would be improved by clearer reporting of (i) the selection criteria for participants and samples included in the analysis, and (ii) previous seropositivity at baseline or potential exposures that might impact subsequent immunogenicity.

Another significant limitation in assessing immunogenicity to Bundibugyo virus is that the antigens used in the assays varied across assays and studies and might not accurately reflect immune responses to the critical strains of Ebola and Bundibugyo virus. For example, the Ervebo vaccine contains the Kikwit strain of Ebola virus and we thus used Kikwit binding responses as our outcome of interest where antibody responses were reported for multiple Ebola virus strains.

However, clinical efficacy of Ervebo was assessed in a cluster randomized trial protecting against the Makona strain^2,33^. Similarly, many studies used the 2007 Uganda strain of Bundibugyo virus, rather than the 2026 strain^19,21,22^. An ideal study would compare binding or neutralization to the Ebola virus Makona antigen (where efficacy is established^2^) and drop in titer to the Bundibugyo 2026 antigen. However, only 1 of 6 (17%) of the included studies analyzed both the Ebola Makona and the Bundibugyo 2026 antigen^20^. It is not clear whether strain differences will affect estimates, but a previous systematic review and meta-analysis suggests that different viral strains may have a significant impact on antibody responses^34^. Lhomme et al.^22^ who report antibody responses after either Ervebo and Zabdeno/Mvabea vaccination, also report binding to both the Ebola Kikwit and Mayinga glycoprotein with higher Bundibugyo to Ebola fold-differences for Mayinga than Kikwit. Nonetheless, using Mayinga instead of Kikwit as a comparator for the fold-drop meta-analysis resulted in a comparable estimate for the fold-drop from the Ebola virus to the Bundibugyo virus antibody response after Ervebo vaccination (**Figure S6**, **Figure S8**).

Finally, this study pooled results from a variety of different assays. It is not clear that the ‘fold change’ in antibody binding in one assay is comparable with the fold-change across assays used in other studies. Fold-change in antibody recognition is often reported as a change in titer (the dilution of antibody that provides a given effect) or concentration. However, the majority of our studies report antibody binding as either MFIs or AUCs rather than titers or concentrations (**Table 1**). Antibody standards or standard curves (to demonstrate how the measured response changes with dilution) would facilitate comparisons but were not presented in the included studies. The fold-change in binding as an MFI or AUC may not scale linearly with, for example, an endpoint titer if binding is very high or low within the detection range of the assay. Of note, the aggregate central estimate of this fold-drop is similar to the studies by Hoffmann et al.^20^ and Ehrhardt et al.^18^ where endpoint titers or NT50s were available or could be computed from the available data.

Our preliminary analysis of cross-reactivity of immune responses to Ervebo focused on only one aspect of immunity – serum antibody binding and neutralization. Other aspects of immunity such as cellular immune responses may also play a role. However, at present there are no reports of T cell responses to BVD after either vaccination or infection.

In addition to studying Bundibugyo virus cross reactivity after vaccination in humans, we also analyzed clinical protection after rVSV-EBOV vaccination in animal models of Bundibugyo virus infection. We identified only two studies reporting survival after vaccination with an rVSV-based vaccine and Bundibugyo virus challenge in an animal model. Importantly, to the best of our knowledge this is all the currently available data that meets our inclusions and exclusion criteria. Encouragingly, after comparing our included studies with studies identified discussed in the WHO Interim guidance (31 August 2026)^35^ and an Africa CDC announcement^32^ we found that we had considered all the studies identified in this guidance with the exception of unpublished data (e.g. Lehrer et al., 2026 is an unpublished study cited in the WHO Interim guidance). While these studies indicated a trend for protection against death, the available evidence from animal models is very limited, with only two studies both of which are at high risk of bias, and a total of 24 animals (8 non-human primates and 16 ferrets). We restricted our analysis to animal studies that reported rVSV-based vaccination with Ebola virus antigens only. This was to allow for inclusion of evidence from studies that used a research-grade Ervebo vaccine. i.e. laboratory constructs that mimic Ervebo rather than being the licensed vaccine. However, effectiveness of an rVSV-based vaccine may not accurately reflect the effectiveness of the Ervebo vaccine, which may differ in the vaccine dosage and stability or other properties. Moreover, the restriction of including only vaccines with Ebola virus antigen led to the exclusion of a study by Mire et al.^36^ that reported survival after Bundibugyo challenge in cynomolgus macaques vaccinated with vaccines containing either a Bundibugyo virus antigen or a combination of Ebola virus antigens and Sudan virus antigens (combined or as prime-boost regimen). Mire et al.^36^ reported that 2 of 3 nonhuman primates died after Bundibugyo challenge after single injection with a vaccine containing both Ebola and Sudan virus antigens (2 of 3 control animals died, RR: 1.00, 95% CI: 0.28 – 3.64). In the same study, a prime-boost vaccination (Sudan virus antigen followed by Ebola virus antigen booster 14 days later) protected 3 of 3 animals (RR: 0, 95% CI: 0 - 1.25). Thus, this indicates potential for protection after prime-boost vaccination but was not statistically significant. Further evidence of Ervebo protection in animal studies would be helpful in assessing vaccine protection. More detailed reporting on randomization methods and inclusion of a comparable baseline group in these studies could improve the risk of bias assessment in future studies.

In the absence of clinical efficacy studies, data on antibody cross reactivity in humans and protection in animal models has been invoked in discussions on the potential deployment of Ervebo in the BVD emergency (along with observational data on health care worker infections)^32^, and 50,000 doses of Ervebo have been provided for protection of healthcare workers^37^. This work presents, to the best of our knowledge, the first systematic review of Bundibugyo cross-reactivity of the Ervebo vaccine and protective efficacy of rVSV-based Ebola vaccines against Bundibugyo virus challenge in animal studies with an assessment of risk of bias and a meta-analysis of Bundibugyo virus cross-reactivity. Further studies of Ervebo immunogenicity and protection are ongoing, and it is important that new data is incorporated into the meta-analysis as it becomes available.

## Data Availability

All data and code are available upon request to the authors and will be made publicly available upon publication.

## Acknowledgements

We thank the authors of Hoffmann et al. (2026)^20^ and Mdluli et al. (2025)^21^ for providing data from their published studies.

This work is funded by the National Health and Medical Research Council (NHMRC, Australia) Investigator Grants 2034282 (to ES), 2026360 (to DC), 2034108 (to MPD), 2033318 (to DSK).

## Declaration of interests

DSK has provided paid services to a clinical trial unit at QIMR Berghofer testing antimalarial compounds. These included trials sponsored by Merck co, GSK, and Medicine for Malaria Venture. The authors declare no other competing interests.

## Ethics statement

This work was approved under the UNSW Sydney Human Research Ethics Committee (approval HC200242).

## Contributions

Conceptualization: ES, MPD, DSK, DC

Data curation: ES, MC, EE, AM

Formal analysis: ES, KME, TT, MC, EE

Funding acquisition: ES, MPD, DSK, DC

Methodology: TT, ES, DSK, MPD, MC, EE, BAB

Project administration: BAB

Supervision: ES, MPD, DK, TT

Writing – original draft: ES, MPD,

Writing – review & editing: All authors

## Notes

### Author Declarations

The study used only openly available human data that were identified through a publicly available evidence map at: https://livingevidence.org.au/research-initiatives/about-feeva/evidence-maps/.

