## Supplementary Material for "Evidence for cross-reactivity and protection against Bundibugyo virus disease after heterologous vaccination: a systematic review and meta-analysis"

**Supplementary Methods** 3

***Systematic review: Bundibugyo immunogenicity in humans after Ervebo or Zabdeno/Mvabea vaccination*** *3*

*Inclusion and exclusion criteria* 3

***BDBV immunogenicity data analysis*** *3*

*Ehrhardt et al. (2019)* 3

*Halbrook et al. (2026)* 4

*Hoffmann et al. (2026)* 4

*Hood et al. (2026)* 4

*Lhomme et al. (2026)* 5

*Mdluli et al. (2025)* 5

***Systematic review: Animal models of rVSV vaccination and BDBV challenge*** *6*

***Meta-analyses and statistical methods*** *6*

**Supplementary Figures** 7

**Figure S1** PRISMA flowchart for the selection of studies reporting Bundibugyo immunogenicity in humans after Ervebo or Zabdeno/Mvabea vaccination. 7

**Figure S2** PRISMA flowchart for the selection of studies reporting survival in animals challenged with Bundibugyo virus after rVSV-based vaccination (containing Ebola virus antigens only). 8

**Figure S3** Funnel plot for BDBV to EBOV antibody fold-drop analysis by antibody response and vaccine. 9

**Figure S4** Fold-drop in BDBV antibody response compared to EBOV antibody response. 10

**Figure S5** Funnel plot for BDBV to EBOV antibody fold-drop analysis by antibody response and vaccine. 11

**Figure S6** Sensitivity analysis using EBOV Mayinga as a comparator in Lhomme et al. (2026): Fold-drop in BDBV antibody response compared to EBOV antibody response. 12

**Figure S7** Sensitivity analysis using EBOV Mayinga as a comparator in Lhomme et al. (2026): Funnel plot for BDBV to EBOV antibody fold-drop analysis by antibody response and vaccine. 13

**Figure S8** Sensitivity analysis using EBOV Mayinga as a comparator in Lhomme et al. (2026): Fold-drop in BDBV antibody response compared to EBOV antibody response. 14
**Figure S9** Sensitivity analysis using EBOV Mayinga as a comparator in Lhomme et al. (2026): Funnel plot for BDBV to EBOV antibody fold-drop analysis by antibody response and vaccine. 15

**Supplement References** **16**

### Supplementary Methods

#### Systematic review: Bundibugyo immunogenicity in humans after Ervebo or Zabdeno/Mvabea vaccination

Our study built upon and existing systematic search and evidence map of BDBV vaccination and immunity (the BDBV evidence map)[1,2]. To understand BDBV infection and immunity in humans we first screened studies listed under "Population: Human, vaccination". We found no studies reporting infection outcomes after vaccination. Thus, we searched for studies reporting antibody binding or neutralization after vaccination in humans (specifically: Population = human, Intervention = vaccination, Outcomes = [Assay-Antibody-binding OR Assay-Antibody-neutralizing], including all subcategories of these outcomes). We screened studies to identify studies that reported:

i. binding to both EBOV-GP (Ebola virus glycoprotein of any Ebola virus strain) and BDBV-GP (Bundibugyo virus glycoprotein, any strain) or
ii. neutralizing antibody responses to both EBOV (any Ebola virus strain) and BDBV (any Bundibugyo strain)
iii. after vaccination with either Ervebo or Zabdeno/Mvabea.

Studies were screened independently by two authors (MC, ES). Conflicts were resolved by discussion and consensus.

Additionally, we assessed whether studies identified in on-going screening of the literature met our inclusion criteria.

*Inclusion and exclusion criteria*

Our main question was to assess cross reactivity against BDBV after Ervebo vaccination. However, due to limited available data, we decided to extend our criteria to also include the previously licensed Zabdeno/Mvabea vaccine. While this may not directly inform the cross reactivity of Ervebo against BDBV, it may provide insight into potential cross-protection after heterologous vaccination (since Zabdeno/Mvabea targets multiple filoviruses). Other vaccines that were never licensed and may therefore not have established safety profiles, were excluded. In particular, we also excluded studies (or subgroups in included studies) that report immunogenicity after only a single agent of either Zabdeno or Mvabea alone, or non-standard protocols (such as administering Zabdeno / Mvabea in the opposite order).

#### BDBV immunogenicity data analysis

*Ehrhardt et al. (2019)*

Ehrhardt et al.[3] reported serum responses to EBOV GP (Mayinga) and BDBV GP for 6 study participants (Extended Data Fig. 1 from their published paper). We extracted $OD_{415\,nm-695\,nm}$ by dilution for EBOV GP and the $OD_{415\,nm-695\,nm}$ at the lowest serum dilution for BDBV GP, and used these to estimate the fold-drop in binding between EBOV and BDBV. Specifically, for each individual, we used their $OD_{415\,nm-695\,nm}$ for BDBV GP at a 1:20 dilution and interpolated the equivalent EBOV dilution that would have given this OD (using linear interpolation between $\log_{10}$(dilution) and $OD_{415\,nm-695\,nm}$ for EBOV GP). In our analysis, we used the geometric mean of these fold-drops and the 95% confidence interval of the geometric mean (**Figure 3**).

For each participant the $OD_{415\,nm-695\,nm}$ for BDBV GP at the lowest serum dilution was above the $OD_{415\,nm-695\,nm}$ of non-immunized controls and we thus considered all samples to be above background signals and the assay's limit of detection.

*Halbrook et al. (2026)*

Halbrook et al.[4] showed and reported mean MFI (mean fluorescence intensity) and 95% confidence intervals for multiple time points after vaccination and at two different locations (Beni and Mbandaka in the Democratic Republic of the Congo) in Supplemental Figure 1. We extracted data for 21 days and 6 months post vaccination, data for other times post vaccination were not extracted because Halbrook et al. write that that booster vaccinations were administered towards the end of their study[4].

*Hoffmann et al. (2026)*

Hoffmann et al.[5] shared their raw neutralization data upon written request, i.e. NT50s for all 10 individuals and all antigens for days 0, 28, and 180. We calculated geometric mean titers (GMTs) with censoring regression for EBOV-GP and BDBV 2026-GP for days 28 and 180 (day 0 titers were all below the lowest dilution tested). Hoffmann et al. also reported neutralizing antibody responses for BDBV 2007/08-GP and 2012-GP, but we included only BDBV 2026-GP because the 2026-GP is most relevant for the current outbreak and results were consistent across BDBV antigens. For censoring regression, we used the lowest dilution tested and cut-off (10) as the limit of detection of the assay.

The GMTs for EBOV-GP were the same as reported by Hoffmann et al., i.e. 43.0 and 34.1 for days 28 and 180, respectively, since none of the 10 samples had NT50s below the limit of detection. For BDBV2026-GP, GMTs with censoring were 13.4 and 11.2 for days 28 and 180, respectively. The fold-differences were 3.2- and 3.0-fold lower BDBV 2026 NT50s compared to EBOV NT50s.

For 2007/08 BDBV-GP, we estimated GMTs of 13.6 and 11.2 on days 28 and 180 after vaccination, respectively. Thus, fold-differences to EBOV-GP were 3.2- and 3.0-fold lower BDBV than EBOV responses for days 28 and 180. Similarly, for 2012 BDBV-GP, we estimated GMTs of 13.7 and 11.0 on days 28 and 180, respectively. Thus, fold-differences to EBOV-GP were 3.1- and 3.1-fold lower BDBV than EBOV responses for days 28 and 180. Thus, fold-differences to EBOV-GP were very similar for all BDBV strains reported.

*Hood et al. (2026)*

Hood et al.[6] reported IgG binding (as the area under the curve, AUC, of OD by dilution) to both Ebola virus (EBOV) glycoprotein and BDBDV glycoprotein as well as fold-differences (with IQR) between EBOV and BDBV antibody binding responses for Ervebo vaccinees and EVD survivors. We used the provided fold-difference in our analysis. For the uncertainty in the fold-difference, we calculated the variance of $\log_{10}$-fold-differences from the provided IQR by assuming a normal distribution of $\log_{10}$-fold-differences (using the formula in (Eq. S1). For consistent reporting of the uncertainty in the fold-difference estimate in the different studies, we used the variance to calculate the 95% confidence interval and reported this confidence interval rather than the IQR in the forest plot of Bundibugyo immunogenicity studies (**Figure 3**).

*Lhomme et al. (2026)*

Lhomme et al.[7] reported the antibody response at day 28 and month 3 in Table S2. We extracted the media, Q1 (25$^{th}$ percentile), and Q3 (75$^{th}$ percentile) antibody responses for EBOV Kikwit and Mayinga and BDBV. Medians were used directly to calculate the fold-difference in the BDBV and EBOV antibody response.

We calculated 95% confidence intervals for the fold-difference by assuming $\log_{10}$-antibody responses follow a normal distribution. Thus, we could use reported quantiles to calculate the standard deviation for EBOV and BDBV antibody responses:

$$\sigma = \frac{\log_{10}(\mathrm{Q3}) - \log_{10}(\mathrm{Q1})}{\Phi^{-1}(0.75) - \Phi^{-1}(0.25)} \quad \text{(Eq. S1)}$$

where σ denotes the standard deviation, $\log_{10}(\mathrm{Q1})$ and $\log_{10}(\mathrm{Q3})$ are the $\log_{10}$-quartiles, and $\Phi^{-1}(0.25)$ and $\Phi^{-1}(0.75)$ are the inverse cumulative distribution function of the standard normal distribution evaluated at 0.25 and 0.75, respectively. The variance of $\log_{10}$-antibody responses for EBOV and BDBV are then $\sigma^2_{\mathrm{EBOV}}$ and $\sigma^2_{\mathrm{BDBV}}$, respectively, and the variance of the $\log_{10}$-fold-difference is $\sigma^2 = \sigma^2_{\mathrm{EBOV}} + \sigma^2_{\mathrm{BDBV}}$.

In the main analysis, we included antibody binding to EBOV Kikwit as the comparator for binding to BDBV. Binding to EBOV Mayinga was used in a sensitivity analysis that resulted in very similar estimates for the fold-drop after Ervebo vaccination (**Figure S6** to **Figure S9**).

*Mdluli et al. (2025)*

We analyzed the supplementary data file provided by Mdluli et al.[8] to calculate Geometric Mean (GM) antibody responses. We excluded people with HIV and included only treatment "Ad26, MVA" (Zabdeno/Mvabea) and total IgG for antigens BDBV_Uga07_GP and EBOV_Kik95_GP. This excluded all neutralizing titers since they were only reported for treatment "MVA, Ad26".

Remaining binding antibody levels (reported as adjusted mean fluorescence intensity, MFI) were used to calculate GM MFIs. Specifically, we used censored regression of $\log_{10}$(adjusted MFIs) with limit of detection 200 to calculate GM MFIs for each antigen at each visit.

We found that both EBOV Kikwit and BDBV (Uganda 2007) GM MFIs were above the limit of detection 50 days after vaccination. Geometric mean adjusted MFIs were 14403 for EBOV (57/57, 100% of samples had MFIs > 200) and 338 for BDBV (37/57, 65% of samples had MFIs > 200). Thus, the fold-difference between BDBV and EBOV binding antibody responses 50 days after

Zabdeno/Mvabea vaccination was 42.6-fold lower BDBV binding antibody response compared to the EBOV Kikwit binding antibody response.

**Systematic review: Animal models of rVSV vaccination and BDBV challenge**

We searched the evidence map of BDBV vaccination and immunity (the BDBV evidence map) to identify studies that report:

i. Survival after BDBV challenge of animals that
ii. Received an rVSV-EBOV vaccine (containing only Ebola virus antigens) and unvaccinated control animals.

Studies were screened by two authors (MC, ES) and data were extracted by one author (ES) and verified by another (AM).

**Meta-analyses and statistical methods**

To estimate geometric means of outcomes with censoring (e.g. samples at the limit of detection or lower limit of quantification of an assay), we $\log_{10}$-transformed the outcomes and used censored regression to estimate the mean and variance of the mean (censReg package[9]). If samples did not include censored samples, we directly computed the geometric mean with variance.

For all statistical analyses, a significance level of 0.05 was used, i.e. p-value <0.05. All confidence intervals are 95% confidence intervals, and all statistical analyses were performed using R (version 4.5.2)[10].

### Supplementary Figures

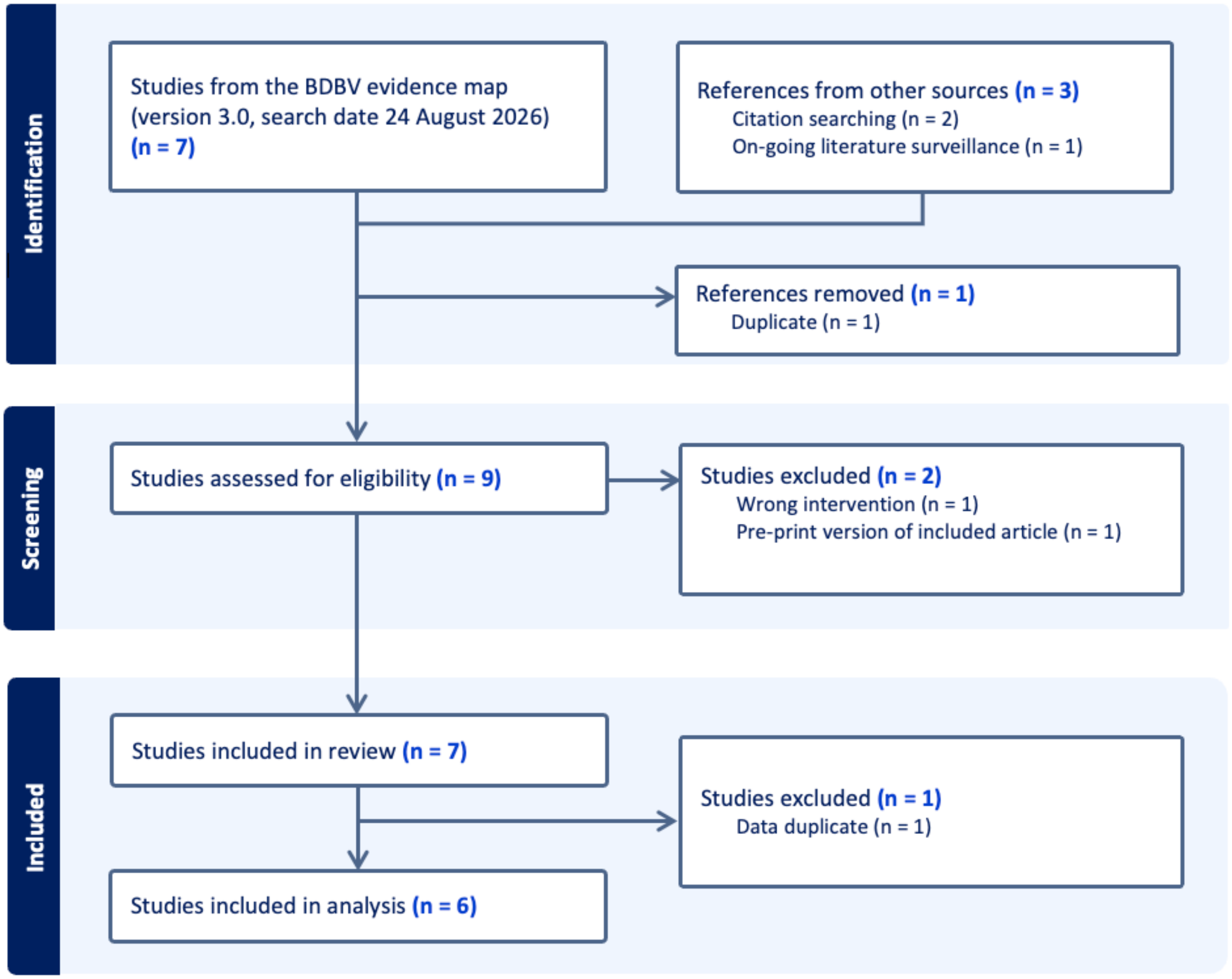


**Figure S1** PRISMA flowchart for the selection of studies reporting Bundibugyo immunogenicity in humans after Ervebo or Zabdeno/Mvabea vaccination.
We search the BDBV evidence map with the following specifications: Population = human, Intervention = vaccination, Outcomes = [Assay-Antibody binding OR Assay-Antibody neutralizing], including all subcategories of these outcomes.

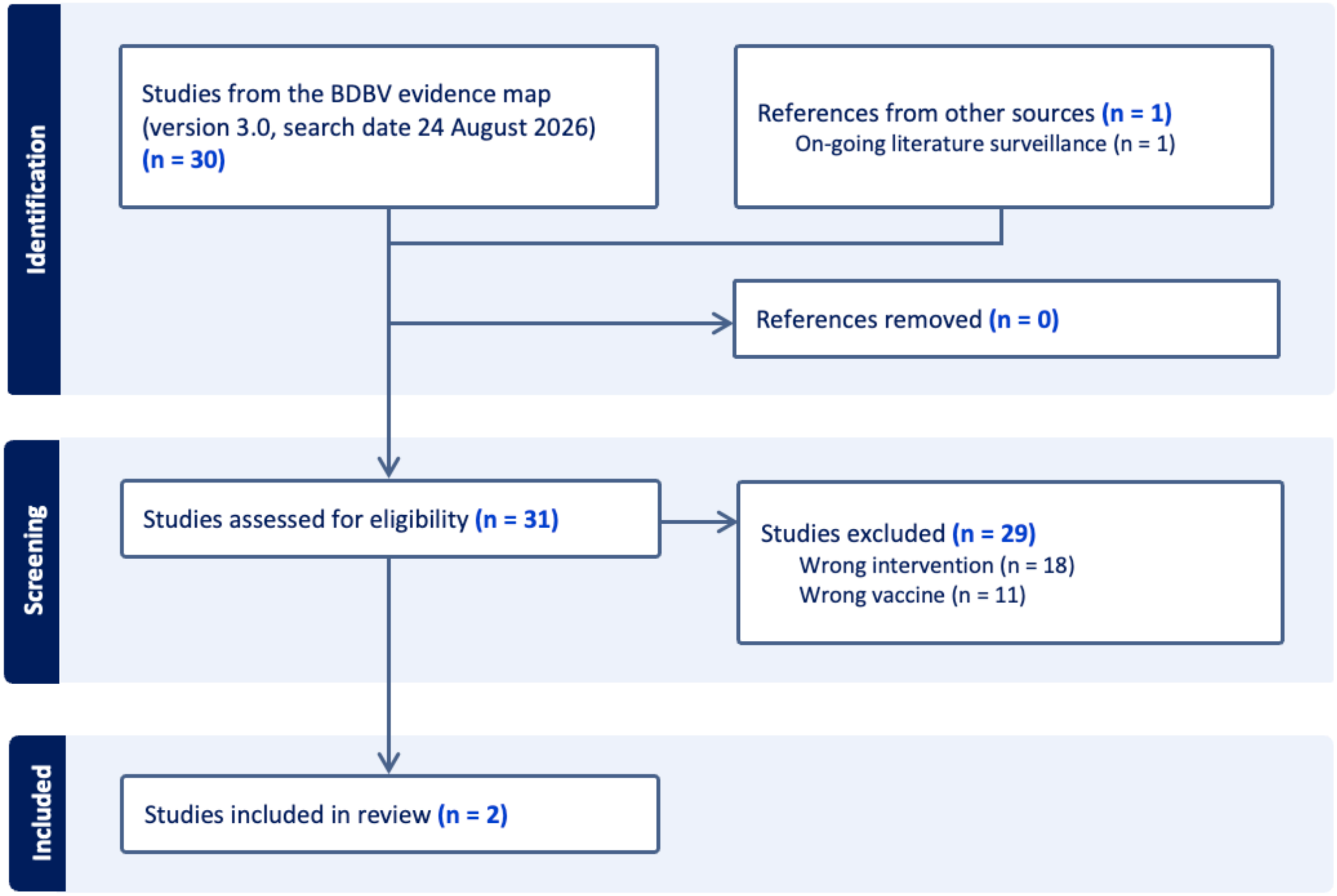


**Figure S2** PRISMA flowchart for the selection of studies reporting survival in animals challenged with Bundibugyo virus after rVSV-based vaccination (containing Ebola virus antigens only).
We searched the BDBV evidence map with the following specifications: Population = [Non-human primate OR mouse OR ferret OR guinea pig OR other animal], intervention = infection challenge, outcome = survival.

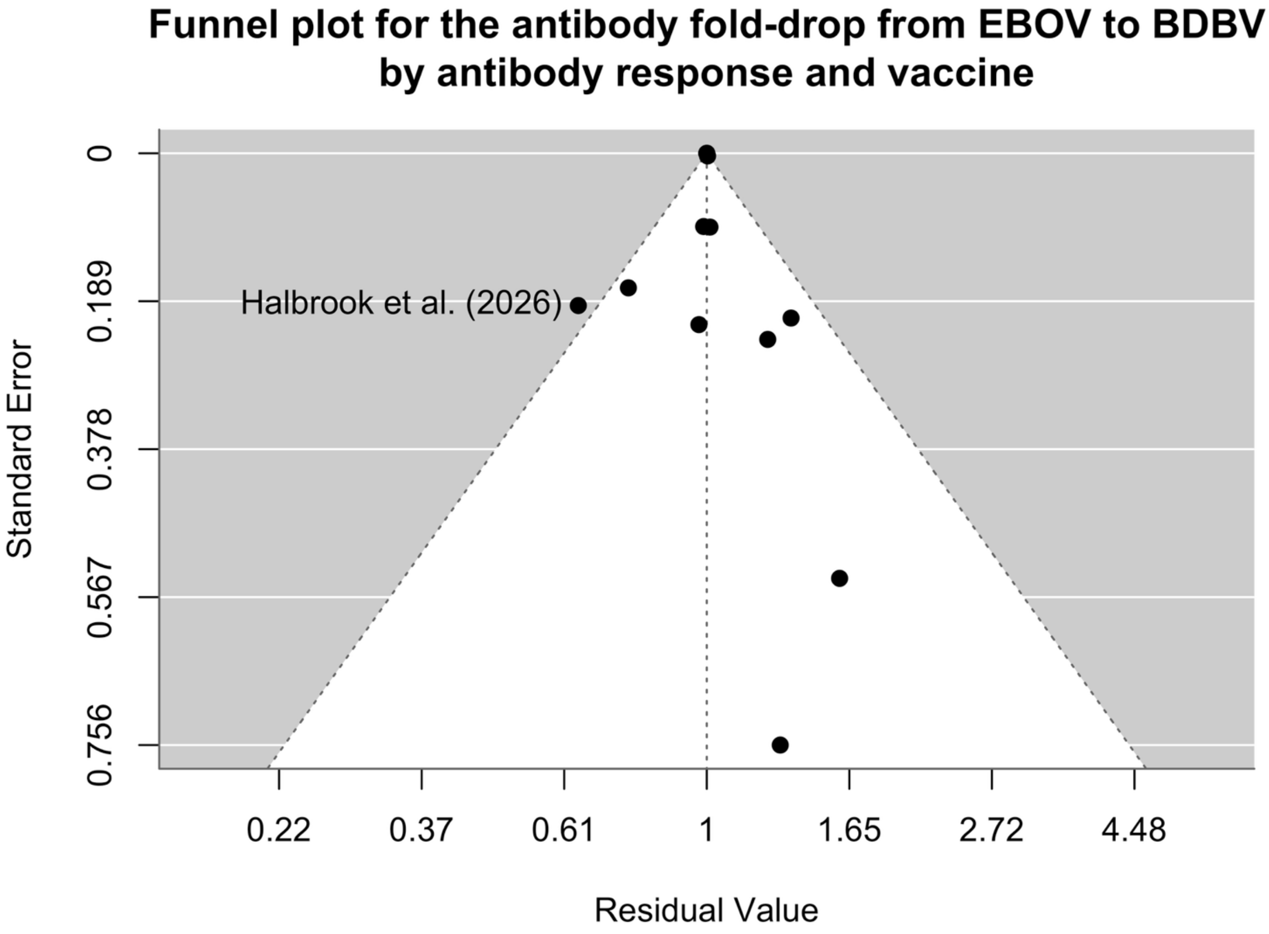


**Figure S3** Funnel plot for BDBV to EBOV antibody fold-drop analysis by antibody response and vaccine.
The associated model output is shown in **Figure 3**. There is one outlier from Halbrook et al. (2026) (Beni, 21 days after vaccination). There is limited power to assess publication bias with so few studies (13 observations from 6 studies).

### Fold–drop from EBOV to BDBV antibody responses

| Study | Vaccine | Day | n | Notes | Fold–drop (95% CI) |
|---|---|---|---|---|---|
| Hoffmann et al. (2026) | Ervebo | 28 | 10 | neut. | 3.20 (1.75–5.83) |
| Hoffmann et al. (2026) | Ervebo | 180 | 10 | neut. | 3.04 (1.67–5.53) |
| Halbrook et al. (2026) | Ervebo | 21 | 599 | Beni | 0.99 (0.41–2.39) |
| Halbrook et al. (2026) | Ervebo | 21 | 482 | Mbandaka | 4.57 (1.56–13.39) |
| Hood et al. (2026) | Ervebo | 28 | 40 | | 2.62 (0.97–7.06) |
| Lhomme et al. (2026) | Ervebo | 28 | 38 | | 5.05 (0.17–153.37) |
| Lhomme et al. (2026) | Ervebo | 90 | 44 | | 8.16 (0.70–94.75) |
| Halbrook et al. (2026) | Ervebo | 180 | 599 | Beni | 1.48 (0.68–3.23) |
| Halbrook et al. (2026) | Ervebo | 180 | 482 | Mbandaka | 5.51 (2.13–14.30) |
| Ehrhardt et al. (2019) | Ervebo | 660 | 6 | | 2.81 (2.60–3.04) |
| **Pooled estimate** | **Ervebo** | | | | **2.81 (2.60–3.03)** |
| Lhomme et al. (2026) | Ervebo (boosted, 56–day spacing) | 90 | 25 | | 10.76 (1.09–106.52) |
| Mdluli et al. (2025) | Zabdeno/Mvabea (28–day spacing) | 50 | 57 | | 42.61 (26.32–68.99) |
| Lhomme et al. (2026) | Zabdeno/Mvabea (56–day spacing) | 90 | 55 | | 6.01 (0.49–73.12) |
| **Pooled estimate** | **Zabdeno/Mvabea** | | | | **39.72 (24.75–63.76)** |

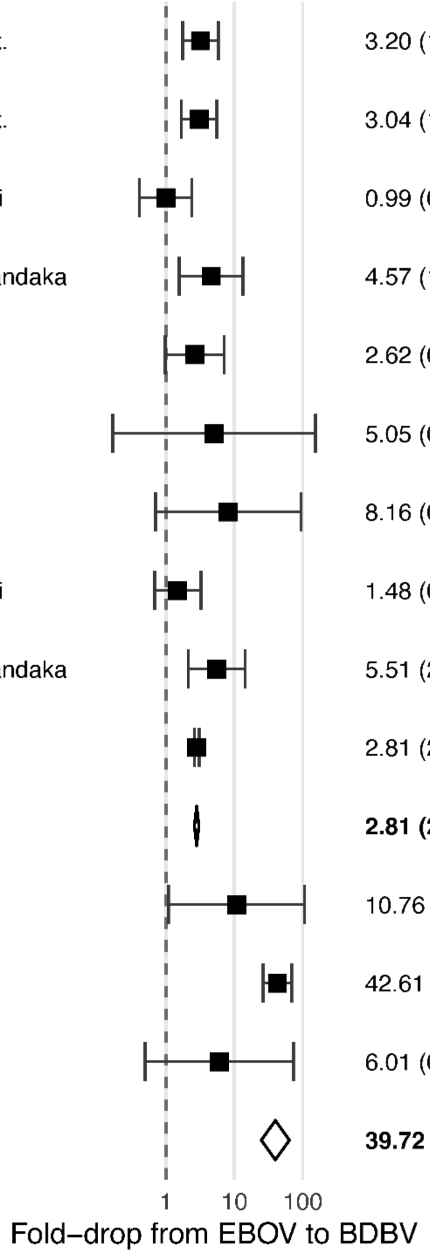


$I^2$: 0.0% (95% CI: 0.0–61.3)
Test for Residual Heterogeneity: QE(df = 11) = 15.38, p = 0.17
Test of Moderators: QM(df = 2) = 940.06, p < 0.0001

**Figure S4** Fold-drop in BDBV antibody response compared to EBOV antibody response.
Data was grouped and a pooled estimated calculated by vaccine used. For pooled estimates, we show 95% prediction intervals rather than confidence intervals (the reported data from each study are confidence intervals). The associated funnel plot is shown in **Figure S5**. Abbreviations: *CI* confidence interval, *n* number of samples, *neut.* neutralizing antibody response.

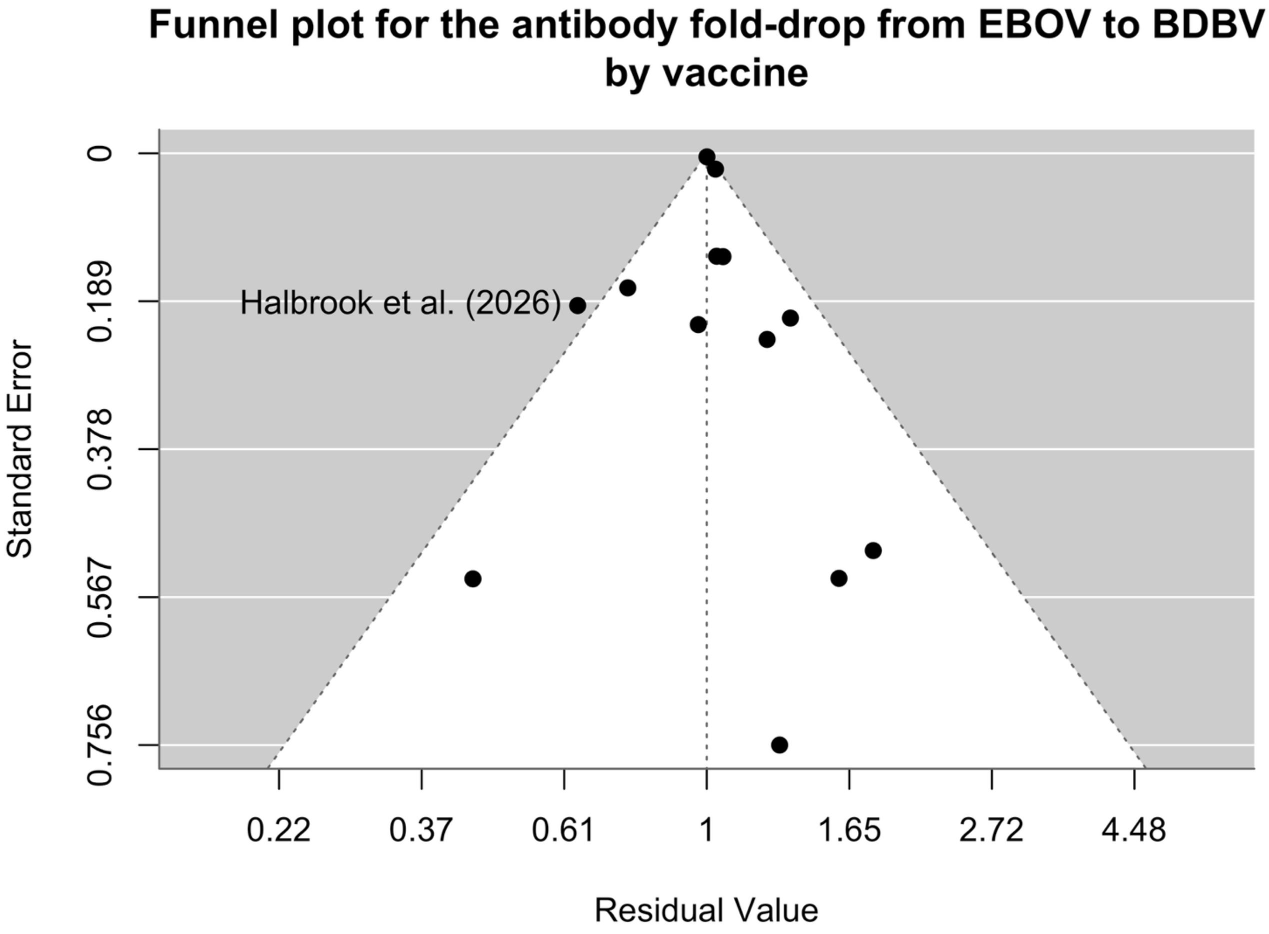


**Figure S5** Funnel plot for BDBV to EBOV antibody fold-drop analysis by antibody response and vaccine.
The associated model output is shown in **Figure S4**. There is one outlier from Halbrook et al. (2026) (Beni, 21 days after vaccination). There is limited power to assess publication bias with so few studies (13 observations from 6 studies).

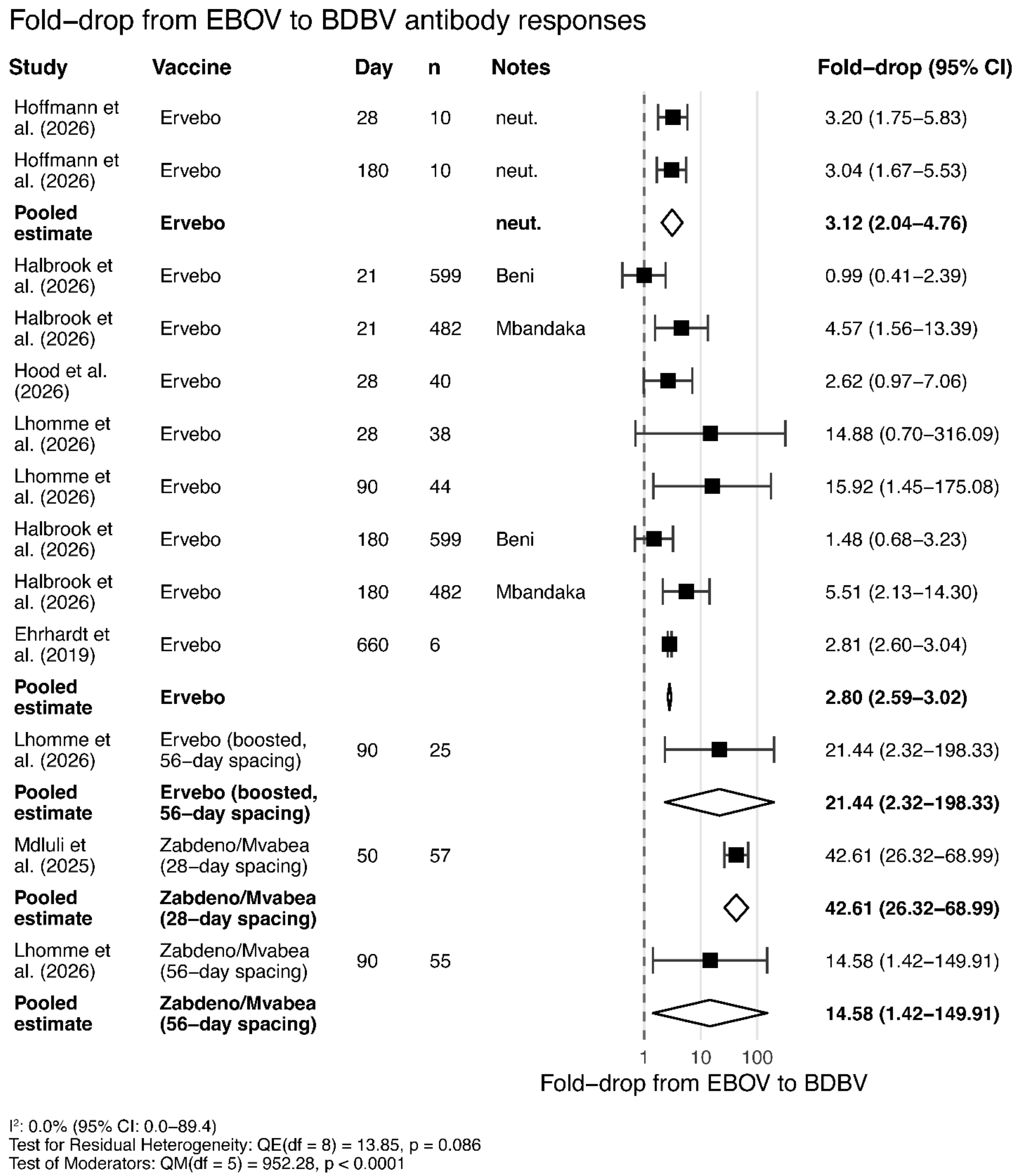


**Figure S6** Sensitivity analysis using EBOV Mayinga as a comparator in Lhomme et al. (2026): Fold-drop in BDBV antibody response compared to EBOV antibody response.

Data was grouped and a pooled estimated calculated by whether neutralizing (indicated by "neut." in the notes column) or binding antibodies (where 'neut.' not indicated) were reported, and according to the vaccine used. Other entries in the notes column indicate the location from which samples were collected (where data from vaccination at different locations were reported in the same paper). For pooled estimates, we show 95% prediction intervals rather than confidence intervals (the reported data from each study are confidence intervals). The associated funnel plot is shown in **Figure S7**. Abbreviations: CI confidence interval, n number of samples, neut. neutralizing antibody response.

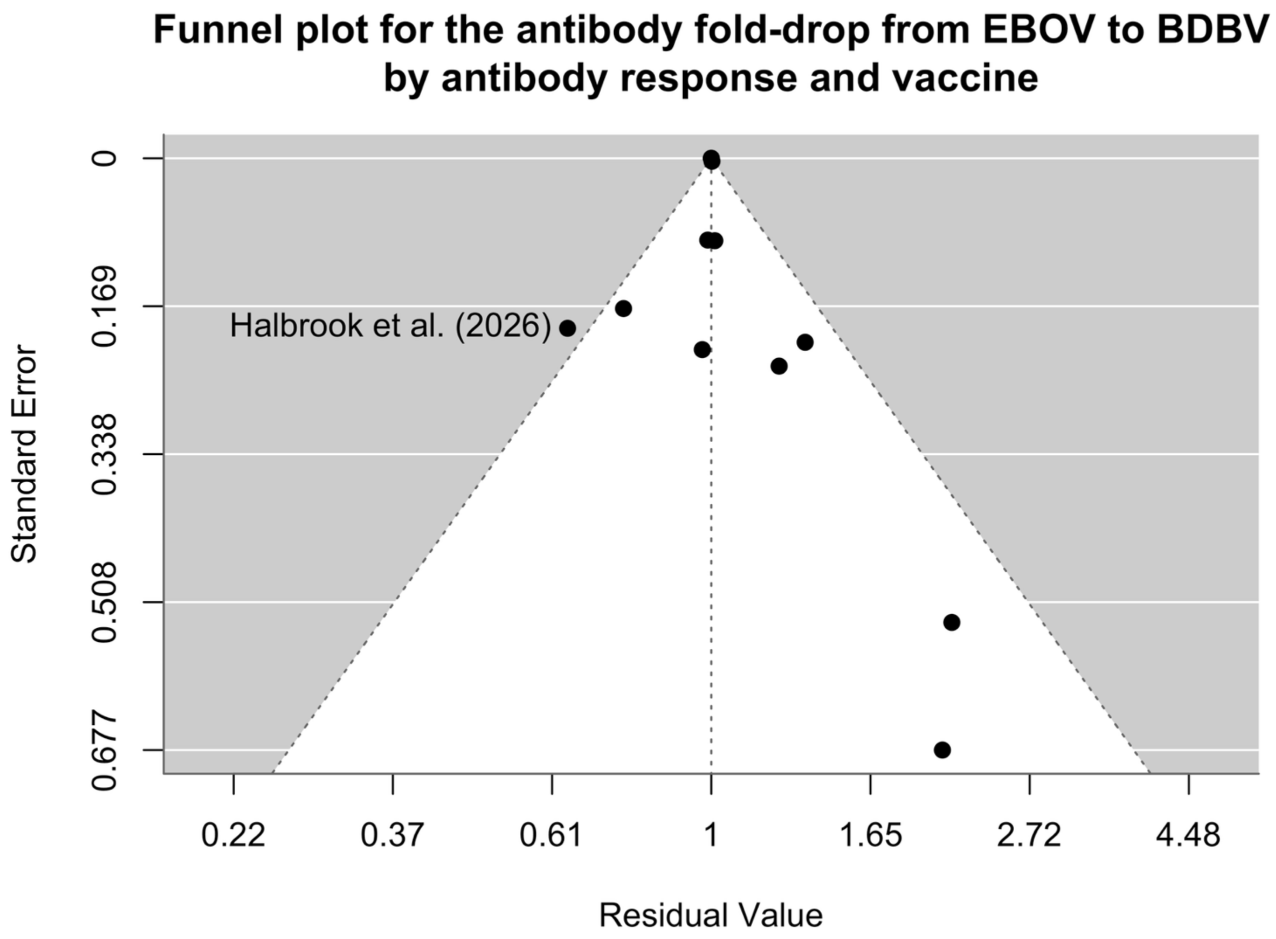


**Figure S7** Sensitivity analysis using EBOV Mayinga as a comparator in Lhomme et al. (2026): Funnel plot for BDBV to EBOV antibody fold-drop analysis by antibody response and vaccine.
The associated model output is shown in **Figure S6**. There is one outlier from Halbrook et al. (2026) (Beni, 21 days after vaccination). There is limited power to assess publication bias with so few studies (13 observations from 6 studies).

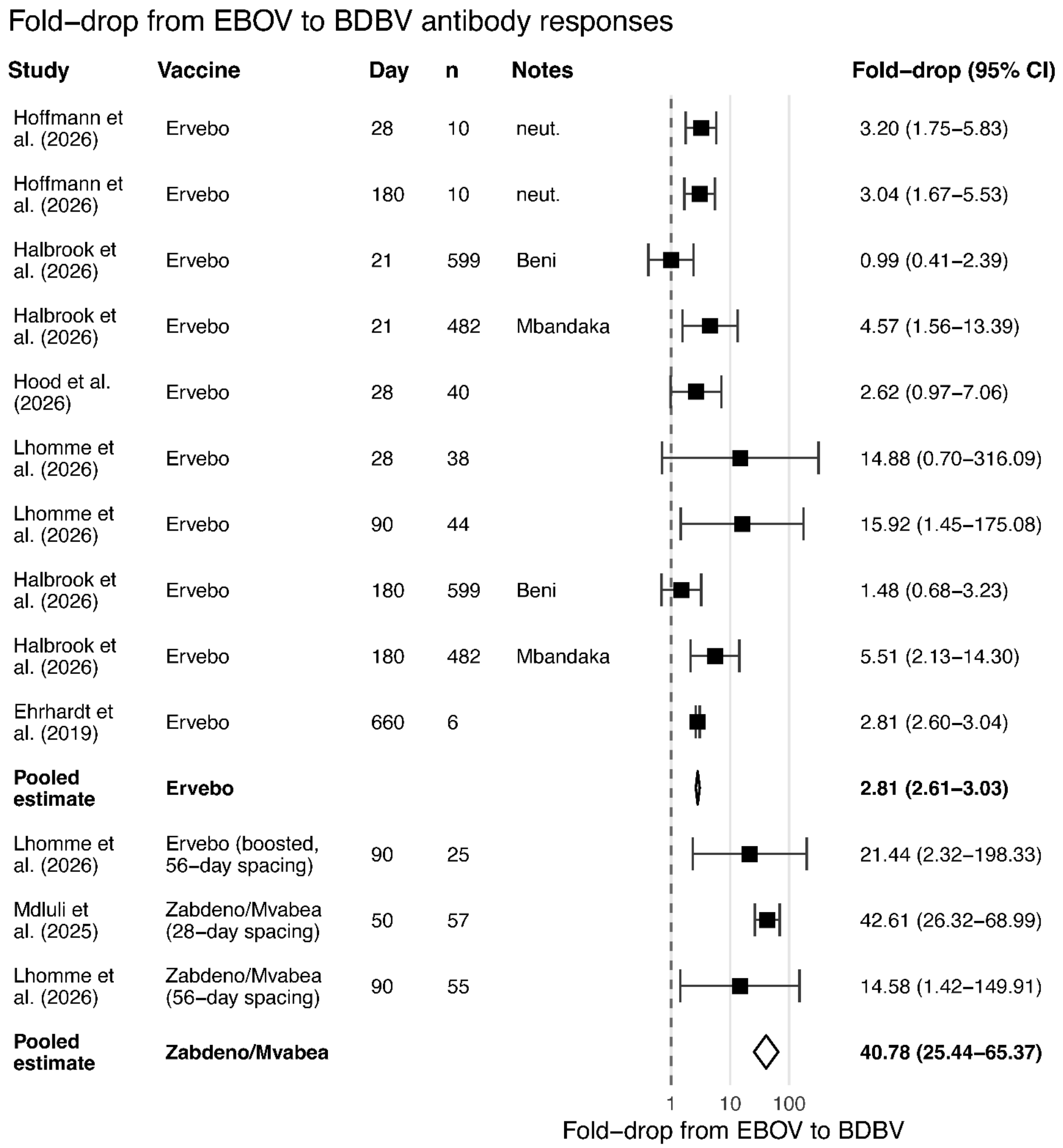


$I^2$: 0.0% (95% CI: 0.0–74.3)
Test for Residual Heterogeneity: QE(df = 11) = 18.08, p = 0.080
Test of Moderators: QM(df = 2) = 948.05, p < 0.0001

**Figure S8** Sensitivity analysis using EBOV Mayinga as a comparator in Lhomme et al. (2026): Fold-drop in BDBV antibody response compared to EBOV antibody response.

Data was grouped and a pooled estimated calculated by vaccine used. For pooled estimates, we show 95% prediction intervals rather than confidence intervals (the reported data from each study are confidence intervals). The associated funnel plot is shown in **Figure S9**. Abbreviations: *CI* confidence interval, *n* number of samples, *neut.* neutralizing antibody response.

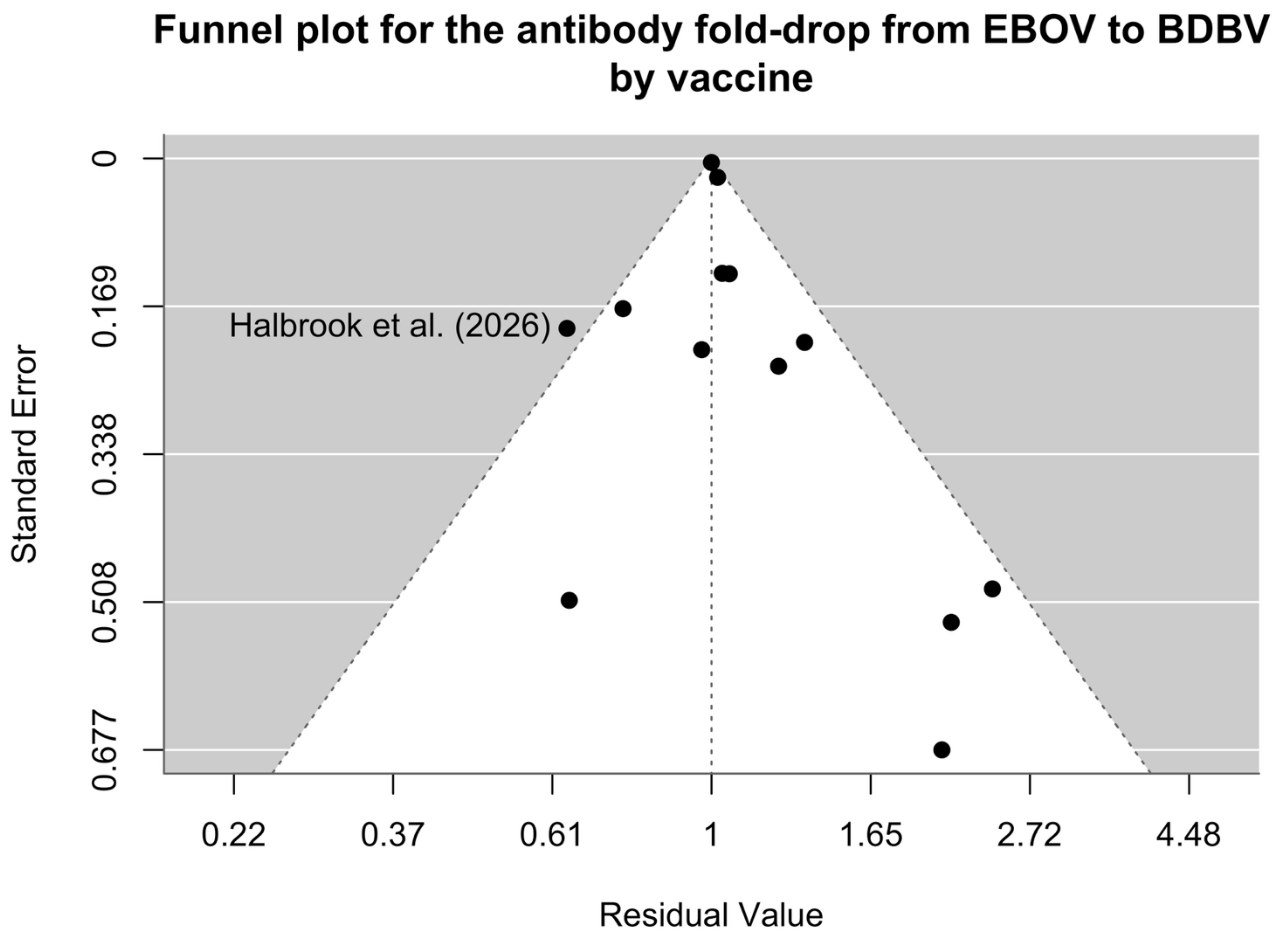


**Figure S9** Sensitivity analysis using EBOV Mayinga as a comparator in Lhomme et al. (2026): Funnel plot for BDBV to EBOV antibody fold-drop analysis by antibody response and vaccine.

The associated model output is shown in **Figure S8**. There is one outlier from Halbrook et al. (2026) (Beni, 21 days after vaccination). There is limited power to assess publication bias with so few studies (13 observations from 6 studies).
